# Identification of Comprehensive Genetic Factors, Pathways, and Shared Genetic Architecture of Putamen Volume in Adolescent Cohort

**DOI:** 10.1101/2025.09.24.25336566

**Authors:** Abanish Singh, Jonathan Posner

**Author notes:** Correspondence: Abanish Singh, Ph.D. Duke University School of Medicine Duke Box 102506, Durham, NC 27705.

## Abstract

The putamen plays a key role in motor control, learning, and cognition, with abnormal putamen volumes associated with neuropsychiatric disorders. Using data from the ABCD study, we performed a genome wide association study (GWAS) of putamen volumes, followed by replication and pathway enrichment analyses. We next evaluated the shared genetic architecture of putamen volume and neuropsychiatric disorders—including depression, schizophrenia, Parkinson’s, ADHD, bipolar, and OCD— using SNP associations from this and prior GWASs. We identified 199 genome-wide significant SNP associations in White participants. Most identified SNPs were in gene regulatory regions and in the neuronal growth-linked genes, *DCC* and *DSCAM*. Sixteen of the most significant associations observed in Whites were replicated in non-White participants. Twenty-one SNPs from prior GWASs of putamen volumes were also replicated in our ABCD GWAS analysis, including five of the top eight SNPs. There was considerable genetic heterogeneity between White and non-White participants in putamen-linked SNPs with significant differences between the minor allele frequencies across the two groups (Wilcoxon rank-sum test Exact prob < 0.0001). We identified a key pathway (*REACTOME_DSCAM_INTERACTIONS)* associated with putamen volume that involves *DSCAM* gene, netrin-1 protein and/or *DCC* gene. In addition, 28 unique SNPs from prior GWASs of neuropsychiatric disorders were strongly associated with putamen volume at Bonferroni-corrected significance, while 40 SNPs shared by at least three disorders were associated with putamen volume at a 0.05 threshold. Our findings provide deeper insights into the shared genetic architecture and cross-population differences in genetic associations of putamen volume.

## INTRODUCTION

The putamen, a structure in the basal ganglia, is crucial for motor control, learning, and cognitive functions. Abnormal putamen volumes have been associated with several neuropsychiatric disorders ^1^, including Parkinson’s disease ^2^, Huntington’s disease ^3^, schizophrenia ^4^; obsessive-compulsive disorder (OCD) ^5^, bipolar disorder ^6^, and attention impairment in attention deficit hyperactivity disorder (ADHD) ^7^. Despite the potential importance of putamen volumes in neuropsychiatric disorders, its genetic underpinning, particularly in youth, remains poorly characterized.

In several neuropsychiatric and neurodegenerative disorders, including Parkinson’s ^2^, Huntington’s ^3^, Alzheimer’s disease ^8^, ADHD ^7^, and major depression ^9^, putamen volumes are typically reduced. In contrast, enlargement of the putamen has been observed in bipolar disorder ^6^, schizophrenia ^4^, autism ^10^, and OCD ^5^. Dopamine dysregulation and neuroinflammation may give rise to these structural changes in conditions like Schizophrenia and OCD ^11^. In major depression, reductions in putamen volume have been associated with impaired reward sensitivity and motivational deficits. Smaller putamen volumes, for example, correlate with symptoms of anhedonia in major depression patients ^12,13^. The increased putamen volume observed in schizophrenia and bipolar disorder, particularly in patients treated with antipsychotic or mood-stabilizing medications, is believed to result from drug-induced neuroplastic changes, leading to structural enlargements ^14,15^. Genetic predispositions and environmental stressors also contribute to alterations in putamen volume ^16^.

Putamen volume heritability is estimated between 71–79% ^17^, with twin studies confirming genetic influence ^18^. Genome-wide association studies have identified loci, such as *KTN1,* ^17^, *DCC* ^19,20^, *BCL2L1* ^17,20^, linked to putamen volume, suggesting that neural growth and plasticity of the putamen are influenced by genetic variations. There may also be genetic overlap between putamen volume and neurodegenerative diseases like Huntington’s disease ^21,22^ and Parkinson’s disease ^23^, where putamen atrophy is pronounced, and neuropsychiatric diseases like schizophrenia, where dopamine-related genes are associated with the disease ^24,25^. There may also be a shared genetic architecture between putamen volume and “broad” depression, including major depression ^26^ and depressive symptoms.

The aforementioned genetic studies were not specifically focused on adolescent populations. The genetic underpinning of putamen volume may differ depending on the age of participants, due to volumetric variation with age ^27^. Changes in putamen volume in older individuals may not occur for the same reasons as they do in the younger populations. Moreover, many neuropsychiatric disorders emerge during adolescence (including ones associated with altered putamen volume). Thus, studying the relationship between genetic variants and putamen volume among youth may be particularly important to gain insights into disease course and etiology. Using the Adolescent Brain and Cognitive Development (ABCD) dataset, we performed a main-effect GWAS of putamen volume, followed by replication analyses and pathways enrichment scoring analysis. Second, we examined whether candidate single nucleotide polymorphisms (SNPs) identified in prior GWASs of putamen volumes would replicate in the ABCD dataset. Third, we examined whether SNPs associated with neuropsychiatric disorders – depression, schizophrenia, Parkinson’s disease, ADHD, bipolar disorder, and OCD -- from prior GWASs would associate with putamen volume in ABCD dataset and identified their shared genetic architecture. We hypothesized that our GWAS analysis of the ABCD dataset cohort would (i) identify genetic variants associated with putamen volume (“new SNPs”), (ii) that genetic variants associated with putamen volumes from prior GWASs would replicate in the ABCD dataset (“replicated candidate SNPs”), and (iii) that genetic variants associated with neuropsychiatric disorders from prior GWASs would associate with putamen volume in the ABCD dataset and these genetic variants would share across the neuropsychiatric disorders (“shared genetic architecture”). Lastly, we explored the overlap between new SNPs, replicated candidate SNPs, and SNPs associated with neuropsychiatric disorders from prior GWASs.

## MATERIAL AND METHODS

### Dataset

We used phenotypic, genetic and brain imaging (MRI) data from the Adolescent Brain Cognitive Development (ABCD) Study. ABCD is a large-scale, longitudinal research project aimed at understanding brain development and its impact on cognitive, social, emotional, and academic outcomes in adolescents. Launched in 2016, it follows over 11,000 children across the U.S., beginning at ages 9-10, to track their development into young adulthood. The study employs neuroimaging, behavioral assessments, and biospecimen analyses to explore the influence of genetic, environmental, and lifestyle factors on the developing brain to understand adolescent brain maturation and mental health trajectories ^28,29^. While the study population comprised mostly White and Black participants, there were also participants from other self-declared racial/ethnic groups, including Natives and Islanders (Native American, Alaska Native, Native Hawaiian, Guamanian, Samoan, and Other Pacific Islander), and Asians and Other Races (Chinese, Japanese, Korean, Filipino, Vietnamese, Other Asian, Asian Indians, and Other Races) (Supplemental Figure 1). In this study, we partitioned the data into two groups i.e., White and non-White participants, and used the data from White participants, which was the largest group, for the *<u>discovery</u>* GWAS analysis, and the data from non-Whites participants for the *<u>replication</u>* analysis.

### Genotypic Dataset

We obtained imputed genotyping data for single nucleotide polymorphisms (SNPs) from ABCD release 5. To ensure robust results, we performed quality control (QC) analyses separately for White and non-White participants, respectively, discarding all multiallelic variants. We applied stringent QC procedures on the imputed SNP dataset that were designed to filter out low-quality data and involved several criteria ^30^. Specifically, we excluded individuals with more than 5% missing genotype data, and SNPs with more than 5% missing data were similarly removed. We also applied a minimum minor allele frequency (MAF) threshold of 1%, ensuring that only common variants were included in the analysis. SNPs that significantly deviated from Hardy-Weinberg equilibrium (HWE) at a P-value threshold of less than 1e-6 were excluded, as such deviations may indicate genotyping errors. As part of our QC process, we checked for sex misclassification by comparing the genetic sex inferred from the genotype data to the recorded sex, and we assessed the genetic relatedness of individuals based on identity by descent (IBD), excluding any misclassified or related participants. In addition, we estimated principal components (PCs) to account for population structure and potential confounding due to ancestry. The PCs were calculated based on linkage disequilibrium (LD)-pruned genotype SNPs within each racial group and followed standard practice of using scree plots to determine the optimal number of PCs, yielding 4 PCs each in both White and non-White participants (Supplementary Figures 2 and 3). The QC filters, procedures, and population stratification resulted in high-quality genetic data consisting of 8.28 million SNPs from 6,200 White participants and 12.06 million SNPs from 3,130 non-White participants.

### Phenotypic/demographic dataset

The study involved demographic variables age, sex (M/F), and race/ethnicity. The White participants included 2,896 females (46.71%) and 3,304 males (53.29%), while the non-White participants included 1,240 females (39.62 %) and 1,890 males (60.38 %). The mean age of White and non-White participants was 118.85 (SD=7.49) and 118.71 (SD=7.35) months, respectively.

### Imaging dataset

The imaging component of the study was designed by the ABCD Data Analysis and Informatics Center (DAIC) in collaboration with the ABCD Imaging Acquisition Workgroup. Imaging techniques and assessments were carefully chosen, optimized, and standardized across all sites of data collection ^29^. Structural MRI scans were segmented into cortical and subcortical regions, which were then utilized to define individualized putamen regions for each participant. We used the total putamen volume as the main outcome variable. We also included brain imaging data on total brain volume in our analysis to adjust the genetic associations of putamen volume for this variable.

### Statistical Analysis

We assessed the distribution of the outcome variable (total putamen volume) for deviations from normality and determined that no transformation was necessary to achieve approximate normality within each population group. The distributions are shown in Supplementary Figures 4 and 5, which ensures that the assumption of normality for the outcome variable is met for the forthcoming analysis.

### Main-effect Discovery GWAS

We performed a discovery GWAS analysis to evaluate the SNP main effect (ME) for the total putamen volume using 8.28 million SNPs from 6,200 White participants. We modeled these SNP main effect GWAS analyses under an additive model using linear regression, adjusting for age, sex, brain volume, and ancestry PCs to control for population structure within each racial group, as shown below:

### Total Putamen Volume ∼ SNP + Age + Sex + Brain Volume + ancestry PCs

The genome-wide significance of ME associations was evaluated at the Bonferroni correction, i.e., 6.04e-9 for White participants.

### Replication of GWAS Associations

Within the non-White sample, we examined the SNPs that showed Bonferroni-corrected significant associations with putamen volume in the discovery GWAS analysis conducted on White participants. For each SNP identified the discovery GWAS analysis, we evaluated the same linear regression association model in non-White participants, i.e., *Total Putamen Volume ∼ SNP + Age + Sex + Brain Volume + ancestry PCs,* and determined the association p-values at the 0.05 significance level.

### Gene Pathway Scoring Analysis

We used the association results (i.e., p-values) from the discovery GWAS to perform pathway enrichment scoring, evaluating the scores of 1077 predefined pathways from the KEGG, BioCarta, and Reactome databases. Briefly, SNPs were mapped to genes, and the association scores for all genes in a pathway were calculated, as implemented in the Pathway Scoring Algorithm ^31^.

### Replication of Associations from Prior GWAS

We searched the NHGRI GWAS catalog ^32^ for all candidate SNPs that have been associated with putamen volume thus far and evaluated their associations in the ABCD dataset for White and non-White participants separately using the aforementioned linear regression model on total putamen volume adjusted by age, sex, brain volume, and the ancestry PCs for each resulting SNP. We considered the replication statistically significant if the p-values were at the 0.05 level in at least one population group, i.e., in White, non-White, or both participants.

### Shared Genetic Architecture of Putamen Volume and Related Psychiatric Disorders

Given that abnormal putamen volume has been linked to depression ^9,26^, schizophrenia ^4^; Parkinson’s disease ^2^, ADHD ^7^, bipolar disorder ^6^, and OCD ^5^, we searched for SNPs associated with these neuropsychiatric disorders and evaluated whether these SNPS were also associated with total putamen volume in the ABCD dataset. We examined this for White and non-White participants separately using linear regression, adjusting for age, sex, brain volume, and ancestry PCs, in a similar manner. The significance of associations for candidate SNPs was evaluated both at a stringent Bonferroni-corrected significance level for SNPs related to each neuropsychiatric trait and at the 0.05 significance level in at least one population group (i.e., White, non-White, or both). We performed analyses and organized the outcomes in two ways to test: 1) whether a SNP associated with a neuropsychiatric disorder, as identified from the NHGRI GWAS catalog, was also associated with putamen volume at a stringent Bonferroni-corrected threshold, and 2) whether a SNP associated with two or more neuropsychiatric disorders, as identified from the NHGRI GWAS catalog, was also associated with putamen volume at a threshold level of 0.05 in White and/or non-White participants in the ABCD dataset.

Lastly, we explored the overlap among SNPs identified in our discovery putamen GWAS, replicated candidate SNPs from prior putamen GWASs, and SNPs associated with neuropsychiatric disorders from previous GWASs. All data processing and analyses were performed using STATA SE 17.0 (StataCorp LP, College Station, TX, USA), PLINK v1.9 ^33^, or R version 4.3.1 with RStudio, 2023.06.1 Build 524, © 2009-2023.

## RESULTS

### Main-effect Discovery GWAS

Figures 1 A and B display the Manhattan and QQ plots of SNP p-values from the GWAS analyses of total putamen volume among White participants in the ABCD study. As depicted in these plots, we observed very strong GWAS signals on chromosomes 14, 18, and 21, which included 199 genome-wide significant SNP associations. A list of the top genome-wide SNPs from White participants is shown in Table 1. The pie chart showing the proportions of functional contexts for all genome-wide significant SNPs is presented in Figure 1 C. Of these, 169 and 13 intronic variants were from the protein-coding genes *DCC* (chromosome 18) and *DSCAM* (chromosome 21), respectively. Seven SNPs were from the long non-coding RNA (lncRNA) *LOC124903320* (chromosome 14). Among the most significant SNPs, five (rs8017172, rs1953350, rs1953351, rs1959089, and rs1953352) were regulatory region variants, located in enhancer regions and CCCTC-binding factor (CTCF) protein binding sites. All genome-wide significant SNPs from the *DSCAM* gene were in strong linkage disequilibrium (LD) (R² = 0.848–1). The SNPs from the *LOC124903320* gene were also in LD with R² = 0.76–1. The SNPs from the *DCC* gene showed strong LD in multiple groups, covering all SNPs with R² = 0.80–1. While *LOC124903320* gene is an uncharacterized locus (https://www.ncbi.nlm.nih.gov/gene/124903320), published literature suggests that both *DCC* and *DSCAM* have been linked with neuronal growth and human central and peripheral nervous system development ^34–40^.

**FIGURE 1.**
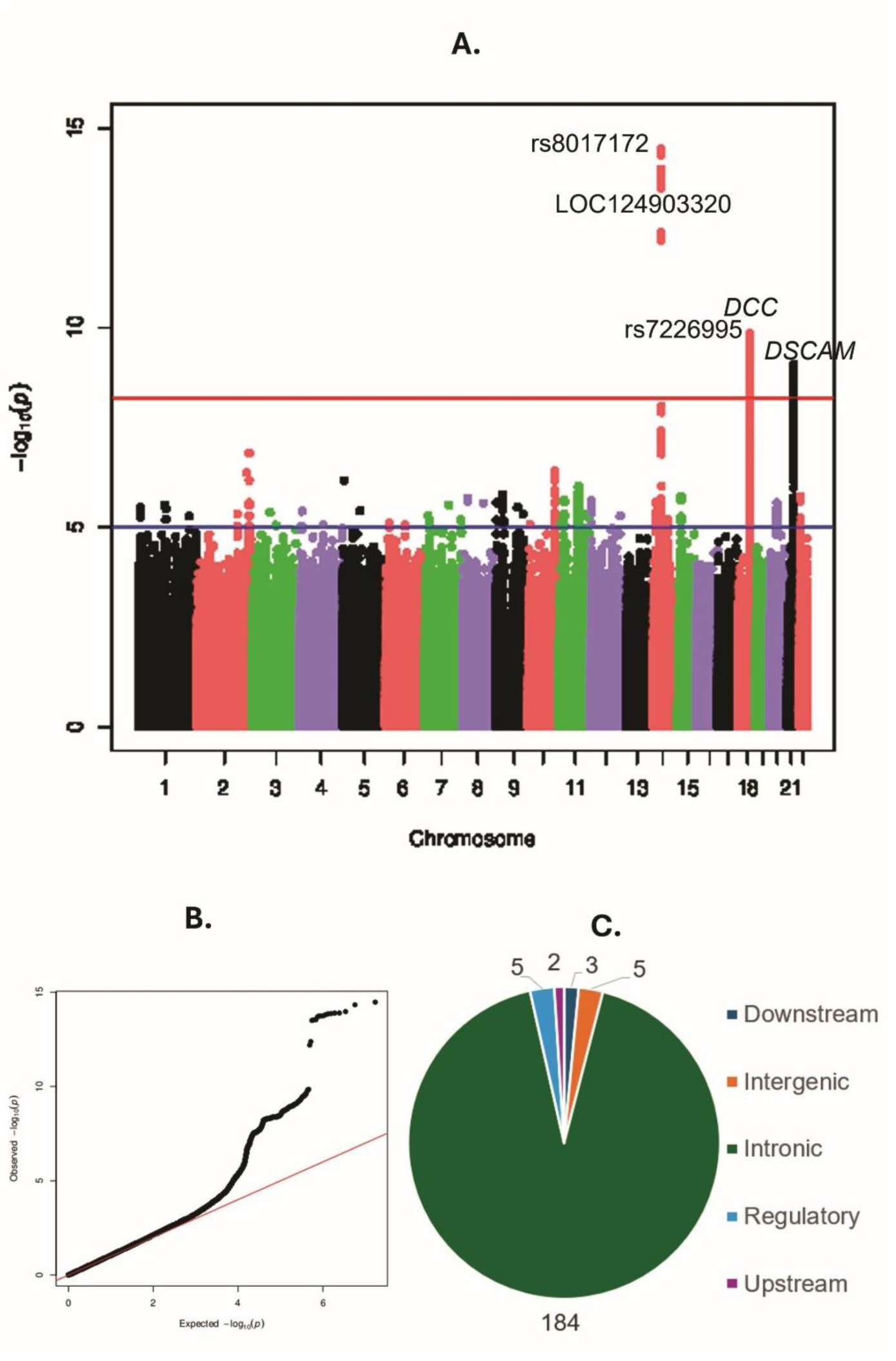
A: Manhattan plot of genome-wide main-effect associations in White participants. B: QQ plot of genome-wide main-effect associations in White participants. C: Pie-chart showing the functional context of discovery GWAS SNPs.

**TABLE 1:** Genome-wide main-effect associations in White participants. Chromosomal base position corresponds to the human reference genome assembly GRCh38.

|  | SNP | CHR | POSITION | BIOTYPE | GENE | CONTEXT | A1<br>WH | MAF<br>WH | A1<br>Non<br>WH | MAF<br>NonW<br>H | BETA WH | P WH | BETA<br>NonWH | P NonWH | PRIOR STUDIES<br>TRAITS (PUBMED ID) |
| --- | --- | --- | --- | --- | --- | --- | --- | --- | --- | --- | --- | --- | --- | --- | --- |
| 1 | rs8017172 | 14 | 55732330 | CTCF binding site | - | regulatory region | A | 0.45 | G | 0.30 | -132.30 | 3.36E-15 | 139.20 | 2.84E-07 | Putamen Vol. (29147026)<br>Parkinson's (28892059) |
| 2 | rs945270 | 14 | 55733755 | - | - | intergenic | G | 0.45 | C | 0.28 | -131.60 | 4.66E-15 | 137.70 | 8.97E-07 | Putamen Vol. (31636452) |
| 3 | rs8014725 | 14 | 55720235 | lncRNA | LOC124903320 | intron | G | 0.47 | A | 0.26 | -130.00 | 1.06E-14 | 151.60 | 7.81E-08 |  |
| 4 | rs10143262 | 14 | 55718343 | lncRNA | LOC124903320 | intron | T | 0.47 | C | 0.27 | -129.70 | 1.26E-14 | 151.30 | 6.53E-08 |  |
| 5 | rs148470213 | 14 | 55726982 | - | - | intergenic | C | 0.49 | T | 0.23 | -129.50 | 1.27E-14 | 144.30 | 2.45E-06 | Putamen Vol. (31636452) |
| 6 | chr14:55722612 | 14 | 55722612 | lncRNA | LOC124903320 | downstream | G | 0.47 | T | 0.26 | -129.50 | 1.36E-14 | 153.20 | 6.67E-08 |  |
| 7 | rs868202 | 14 | 55729044 | - | - | intergenic | C | 0.47 | T | 0.26 | -129.40 | 1.41E-14 | 150.50 | 1.18E-07 | Putamen Vol. (31636452) |
| 8 | rs10129414 | 14 | 55726554 | lncRNA | LOC124903320 | downstream | A | 0.47 | G | 0.28 | -129.20 | 1.57E-14 | 152.50 | 2.74E-08 | Putamen Vol. (31636452) |
| 9 | rs1953350 | 14 | 55730556 | enhancer | - | regulatory region | A | 0.47 | C | 0.28 | -128.90 | 1.82E-14 | 151.00 | 4.00E-08 |  |
| 10 | rs1953351 | 14 | 55730507 | enhancer | - | regulatory region | G | 0.47 | A | 0.28 | -128.90 | 1.82E-14 | 150.80 | 4.16E-08 |  |
| 11 | rs1959089 | 14 | 55731119 | enhancer | - | regulatory region | C | 0.47 | T | 0.28 | -128.60 | 2.05E-14 | 150.80 | 3.92E-08 |  |
| 12 | rs2342588 | 14 | 55730321 | - | - | intergenic | C | 0.47 | T | 0.32 | -127.90 | 2.87E-14 | 147.30 | 2.21E-08 |  |
| 13 | rs2342589 | 14 | 55730350 | - | - | intergenic | A | 0.47 | G | 0.32 | -127.90 | 2.87E-14 | 147.30 | 2.21E-08 |  |
| 14 | rs1953352 | 14 | 55730441 | enhancer | - | regulatory region | C | 0.47 | T | 0.32 | -127.90 | 2.94E-14 | 147.30 | 2.21E-08 |  |
| 15 | rs8012377 | 14 | 55724695 | lncRNA | LOC124903320 | downstream | T | 0.47 | C | 0.28 | -127.80 | 3.14E-14 | 150.30 | 4.02E-08 | Parkinson's (38155330) |
| 16 | rs9671291 | 14 | 55714977 | lncRNA | LOC124903320 | upstream | G | 0.46 |  |  | -121.10 | 4.15E-13 |  |  |  |
| 17 | rs8021018 | 14 | 55710790 | lncRNA | LOC124903320 | upstream | T | 0.45 | C | 0.31 | -120.40 | 6.35E-13 | 91.02 | 5.61E-04 |  |
| 18 | rs7226995 | 18 | 53369385 | protein coding | DCC | intron | A | 0.49 | G | 0.42 | -109.40 | 1.42E-10 | 29.52 | 0.22 |  |
| 19 | rs10502971 | 18 | 53361591 | protein coding | DCC | intron | A | 0.41 | A | 0.28 | 111.10 | 1.92E-10 | 28.73 | 0.28 |  |
| 20 | rs62097996 | 18 | 53316899 | protein coding | DCC | intron | A | 0.41 | A | 0.17 | 110.80 | 2.59E-10 | -2.53 | 0.94 |  |
| 21 | rs62097995 | 18 | 53316863 | protein coding | DCC | intron | A | 0.41 | A | 0.16 | 110.80 | 2.59E-10 | 0.29 | 0.99 |  |
| 22 | rs62098036 | 18 | 53367709 | protein coding | DCC | intron | A | 0.49 | G | 0.49 | -107.20 | 3.20E-10 | 20.50 | 0.39 |  |
| 23 | chr18:53300962 | 18 | 53300962 | protein coding | DCC | intron | A | 0.49 | T | 0.47 | 107.20 | 3.21E-10 | -23.58 | 0.34 |  |
| 24 | rs62097994 | 18 | 53314592 | protein coding | DCC | intron | A | 0.41 | A | 0.17 | 110.00 | 3.40E-10 | -2.05 | 0.95 |  |
| 25 | rs9954838 | 18 | 53312306 | protein coding | DCC | intron | A | 0.49 | C | 0.41 | -107.10 | 3.72E-10 | -6.83 | 0.78 |  |
| 26 | rs11662416 | 18 | 53321414 | protein coding | DCC | intron | G | 0.40 | G | 0.16 | 109.30 | 4.62E-10 | -1.15 | 0.97 |  |
| 27 | rs6508231 | 18 | 53369745 | protein coding | DCC | intron | C | 0.50 | T | 0.41 | -105.80 | 5.49E-10 | 33.65 | 0.16 |  |
| 28 | rs11877400 | 18 | 53368227 | protein coding | DCC | intron | G | 0.50 | T | 0.41 | -105.80 | 5.49E-10 | 32.60 | 0.17 |  |
| 29 | rs60218578 | 18 | 53369246 | protein coding | DCC | intron | T | 0.50 | G | 0.41 | -105.50 | 6.01E-10 | 33.64 | 0.16 |  |
| 30 | rs7227402 | 18 | 53369396 | protein coding | DCC | intron | T | 0.40 | T | 0.26 | 108.20 | 6.18E-10 | 37.49 | 0.16 |  |
| 31 | rs75048819 | 18 | 53318768 | protein coding | DCC | intron | G | 0.40 | G | 0.16 | 108.20 | 6.73E-10 | -0.21 | 1.00 |  |
| 32 | rs9954990 | 18 | 53312275 | protein coding | DCC | intron | A | 0.49 | G | 0.40 | -105.10 | 7.31E-10 | -8.75 | 0.72 |  |
| 33 | rs6508230 | 18 | 53358554 | protein coding | DCC | intron | T | 0.40 | T | 0.26 | 107.30 | 7.71E-10 | 33.62 | 0.21 |  |
| 34 | rs77162730 | 18 | 53318770 | protein coding | DCC | intron | G | 0.40 | G | 0.16 | 107.60 | 8.16E-10 | -0.21 | 1.00 |  |
| 35 | rs2837518 | 21 | 40226239 | protein coding | DSCAM | intron | T | 0.49 |  |  | -101.70 | 8.30E-10 |  |  |  |
| 36 | rs1346914 | 18 | 53319460 | protein coding | DCC | intron | T | 0.40 | T | 0.16 | 107.60 | 8.40E-10 | -3.79 | 0.91 |  |
| 37 | rs8130627 | 21 | 40230912 | protein coding | DSCAM | intron | G | 0.46 |  |  | -101.90 | 8.59E-10 |  |  |  |
| 38 | rs17503448 | 18 | 53331293 | protein coding | DCC | intron | C | 0.40 | C | 0.16 | 107.40 | 9.11E-10 | -4.04 | 0.90 |  |
| 39 | rs77916462 | 18 | 53318766 | protein coding | DCC | intron | G | 0.40 | G | 0.16 | 107.30 | 9.32E-10 | -1.00 | 0.98 |  |
| 40 | rs17504871 | 18 | 53382033 | protein coding | DCC | intron | G | 0.47 | A | 0.28 | -104.80 | 1.02E-09 | -20.90 | 0.44 |  |
| 41 | rs62098023 | 18 | 53367632 | protein coding | DCC | intron | C | 0.40 | C | 0.16 | 107.00 | 1.04E-09 | 7.89 | 0.81 |  |
| 42 | rs2837517 | 21 | 40225992 | protein coding | DSCAM | intron | G | 0.49 |  |  | -101.20 | 1.04E-09 |  |  |  |
| 43 | rs9304444 | 18 | 53312214 | protein coding | DCC | intron | T | 0.49 | G | 0.40 | -103.90 | 1.08E-09 | -8.84 | 0.71 |  |
| 44 | rs11664320 | 18 | 53346253 | protein coding | DCC | intron | C | 0.40 | C | 0.26 | 106.70 | 1.11E-09 | 36.60 | 0.17 |  |
| 45 | rs960697 | 21 | 40232312 | protein coding | DSCAM | intron | G | 0.47 |  |  | -100.90 | 1.11E-09 |  |  |  |
| 46 | rs17412782 | 18 | 53361981 | protein coding | DCC | intron | C | 0.40 | C | 0.16 | 106.50 | 1.14E-09 | 2.55 | 0.94 |  |
| 47 | rs62098022 | 18 | 53362521 | protein coding | DCC | intron | C | 0.40 | C | 0.17 | 106.40 | 1.17E-09 | 1.05 | 0.97 |  |
| 48 | rs62098037 | 18 | 53368355 | protein coding | DCC | intron | C | 0.50 | T | 0.48 | -103.60 | 1.20E-09 | 24.15 | 0.31 |  |
| 49 | rs1504746 | 18 | 53331649 | protein coding | DCC | intron | C | 0.40 | C | 0.16 | 106.60 | 1.20E-09 | 0.48 | 0.99 |  |
| 50 | rs62098014 | 18 | 53342715 | protein coding | DCC | intron | C | 0.40 | C | 0.16 | 106.60 | 1.24E-09 | -1.47 | 0.97 |  |

**TABLE 2:** Gene pathways enrichment scoring analysis from the White participants GWAS analysis.

|  | Name | chi2Pvalue | empPvalue |
| --- | --- | --- | --- |
| 1 | REACTOME_DSCAM_INTERACTIONS | 7.62E-05 | 1.36E-04 |
| 2 | BIOCARTA_BAD_PATHWAY | 2.37E-04 | 3.72E-04 |
| 3 | REACTOME_RECYCLING_PATHWAY_OF_L1 | 8.96E-04 | 1.15E-03 |
| 4 | REACTOME_L1CAM_INTERACTIONS | 2.27E-03 | 2.51E-03 |
| 5 | BIOCARTA_MPR_PATHWAY | 3.43E-03 | 4.65E-03 |
| 6 | BIOCARTA_MYOSIN_PATHWAY | 3.48E-03 | 5.71E-03 |
| 7 | REACTOME_PROCESSING_OF_CAPPED_INTRON_CONTAINING_PRE_MRNA | 3.62E-03 | 3.62E-03 |
| 8 | REACTOME_MRNA_PROCESSING | 3.63E-03 | 3.60E-03 |
| 9 | REACTOME_MRNA_SPLICING | 3.64E-03 | 3.75E-03 |
| 10 | REACTOME_NA_CL_DEPENDENT_NEUROTRANSMITTER_TRANSPORTERS | 3.78E-03 | 5.98E-03 |
| 11 | BIOCARTA_TPO_PATHWAY | 5.31E-03 | 7.62E-03 |
| 12 | BIOCARTA_RANMS_PATHWAY | 5.33E-03 | 3.97E-03 |
| 13 | REACTOME_IL_2_SIGNALING | 7.33E-03 | 9.21E-03 |

### Replication of GWAS Associations

Table 1 also presents the results of aforementioned GWAS SNP associations’ replication in non-White participants. Sixteen of the seventeen most significant SNPs from White participants were also significant in the non-White participants. These SNPs included all five regulatory region variants. However, we observed that the minor allele (A1) was flipped in non-White participants compared to White participants for these sixteen SNPs, indicating genetic heterogeneity between the two population groups. Nonetheless, the direction of the association remained consistent, with the same risk allele being linked with a decrease in putamen volume (see Beta sign in Table 1). In addition, MAF of significant SNPs in White participants (Table 1) varied from 0.40 – 0.50 and varied from 0.16 – 0.49 for the same SNPs in non-White participants. Wilcoxon rank-sum test suggested that the MAFs of the SNPs for the White and non-White participants were not similar and that there was a significant difference between the two paired groups of MAFs (Exact prob < 0.0001).

### Gene Pathway Scoring Analysis

Table 3 presents the outcomes of pathway enrichment scoring analysis for the White participants, where all the most significantly associated pathways are listed. Of these, only one pathway, i.e., *REACTOME_DSCAM_INTERACTIONS* (https://www.reactome.org/content/detail/R-HSA-376172) reached Bonferroni corrected significance (i.e., ∼4.64e-05). The pathway appears to be linked with nervous system development when the *DSCAM* gene binds netrin-1 protein and works alongside or independently of *DCC* gene to guide commissural axons across the midline, cooperatively depending on the cellular and developmental context ^39,41^.

**TABLE 3:**
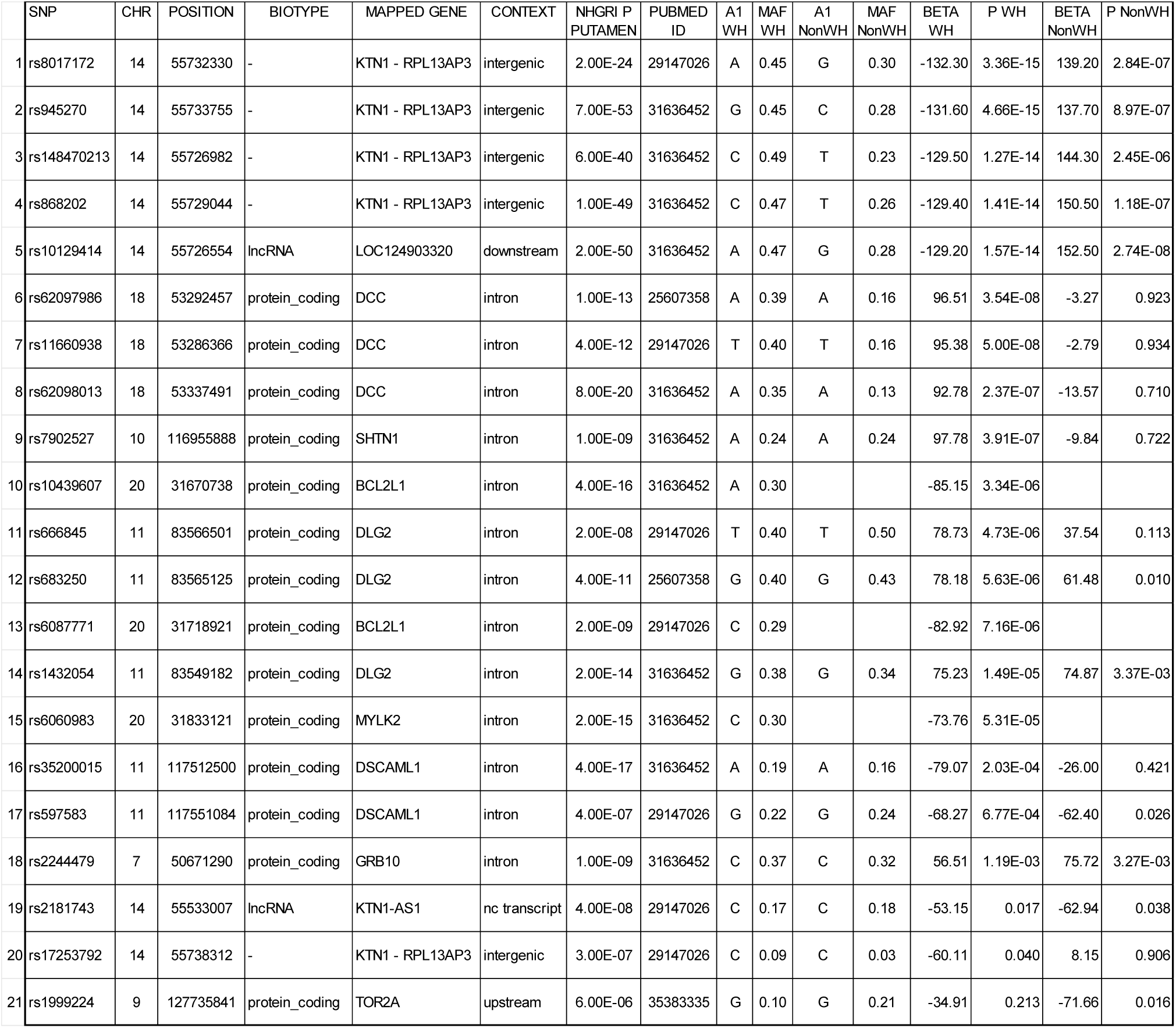
Replication of putamen-associated SNPs from prior studies.

### Replication of Associations from Prior Studies

The NHGRI GWAS catalog ^32^ search revealed that prior GWASs have identified a total of 63 SNPs associated with putamen volume ^17,19,20,42–44^. Of these, 21 SNPs showed significant association with putamen volume in our analysis of White and/or non-White participants, including 14 SNPs that mapped to eight genes (i.e., *DCC*, *DSCAML1*, *BCL2L1*, *MYLK2*, *DLG2*, *GRB10*, *SHTN1*, and *KTN1-AS1*), as presented in Table 3. These outcomes demonstrated the replication of known SNP associations with putamen volume in ABCD participants. Notably, in five SNPs (i.e, rs8017172, rs945270, rs148470213, rs868202, and rs10129414) out of the ten SNPs that displayed a significant association in both White and non-White participants, there was a flip of minor allele, indicating genetic heterogeneity among the putamen-linked SNPs in Whites and non-Whites, consistent with our previous observations; however, there were consistent directions of association with the same risk allele. Finally, the 21 replicated SNPs from prior studies (Table 3) included the five most significant SNPs (i.e., rs8017172, rs945270, rs148470213, rs868202, and rs10129414) from our discovery GWAS list in Table 1, highlighting the robustness of the discovered associations with putamen volume.

### Shared Genetic Architecture of Putamen Volume and Related Psychiatric Disorders

Our NHGRI GWAS catalog ^32^ search identified the following number of SNPs associated with neuropsychiatric disorders in over 300 prior GWASs and other genetic studies: 2,364 SNPs for depression, 4,150 SNPs for schizophrenia, 602 SNPs for Parkinson’s disease, 1,929 SNPs for ADHD, 1,600 SNPs for bipolar disorder, and 275 SNPs for OCD. A subset of these SNPs were available within the quality-controlled ABCD GWAS dataset: 2,030 SNPs for depression, 3,473 SNPs for schizophrenia, 498 SNPs for Parkinson’s disease, 1,744 SNPs for ADHD, 1,344 SNPs for bipolar disorder, and 256 SNPs for OCD.

Table 4 presents the SNPs from prior GWASs of neuropsychiatric disorders that were also associated with putamen volume in the ABCD sample at a stringent Bonferroni-corrected significance threshold. These SNPs included 10 SNPs for depression ^45–49^, 2 SNPs for schizophrenia ^50,51^, 5 SNPs for Parkinson’s disease ^52–56^, 11 SNPs for ADHD ^57–60^, 2 SNPs for bipolar disorder ^48,60^, and 1 SNP for OCD ^61^. Notably, SNPs rs4632195 and rs11663393 from the *DCC*, a gene identified in our discovery GWAS analysis, reached Bonferroni-corrected significance for association with putamen volume in White participants in the ABCD study. In addition to being associated with putamen volume, SNP rs4632195 shared an association with three neuropsychiatric disorders (depression, ADHD, and bipolar disorder) and rs11663393 shared an association with two neuropsychiatric disorders (depression and bipolar disorder). Furthermore, a *DCC* gene SNP rs8084351 from a prior OCD GWAS showed Bonferroni-corrected significant association with putamen volume in White participants. This SNP was also shared in GWASs for depression, schizophrenia, ADHD, and bipolar disorder, as listed in the NHGRI GWAS catalog.

**TABLE 4:** Associations of putamen volume and the SNPs that were implicated with neuropsychiatric disorders in NHGRI GWAS catalog. These candidate SNP associations were assessed at a stringent Bonferroni-corrected significance threshold.

|  | MAPPED TRAIT | SNP | CHR | POSITION | MAPPED GENE | CONTEXT | P NHGRI/<br>GWAS | PUBMED<br>ID | A1<br>WH | A1<br>NonWH | MAF<br>WH | MAF<br>NonWH | BETA<br>WH | P<br>WH | BETA<br>NonWH | P<br>NonWH |
| --- | --- | --- | --- | --- | --- | --- | --- | --- | --- | --- | --- | --- | --- | --- | --- | --- |
| 1 | Depression | rs1431181 | 18 | 53253042 | DCC | intron | 3.00E-12 | 29942085 | A | A | 0.40 | 0.16 | 95.63 | 4.49E-08 | -2.01 | 0.953 |
| 2 | Depression | rs4277413 | 18 | 53172556 | DCC | intron | 1.00E-11 | 29942085 | G | G | 0.47 | 0.29 | 92.09 | 7.11E-08 | -17.04 | 0.522 |
| 3 | Depression | rs17410557 | 18 | 53250021 | DCC | intron | 9.00E-10 | 34586374 | C | C | 0.37 | 0.15 | 93.57 | 1.10E-07 | -6.89 | 0.842 |
| 4 | Depression | rs12968428 | 18 | 53223855 | DCC | intron | 2.00E-12 | 29942085 | G |  | 0.39 |  | -85.81 | 7.79E-07 |  |  |
| 5 | Depression | rs8097041 | 18 | 53371847 | DCC | intron | 2.00E-11 | 29942085 | A | T | 0.39 | 0.47 | -79.97 | 5.00E-06 | 42.18 | 7.97E-02 |
| 6 | Depression | rs7505145 | 18 | 53185406 | DCC | intron | 2.00E-08 | 29942085 | A |  | 0.47 |  | -75.11 | 9.23E-06 |  |  |
| 7 | Depression | rs7231178 | 18 | 53227371 | DCC | intron | 2.00E-09 | 35017462 | G | G | 0.48 | 0.48 | -73.75 | 1.47E-05 | -6.34 | 0.802 |
| 8 | Depression | rs8099160 | 18 | 53226240 | DCC | intron | 2.00E-14 | 29942085 | A | A | 0.44 | 0.27 | 74.02 | 1.51E-05 | -0.18 | 0.995 |
| 9 | Depression | **rs11663393** | 18 | 53088362 | DCC | intron | 2.00E-10 | 31926635 | A | A | 0.45 | 0.32 | 72.85 | 1.72E-05 | 6.26 | 0.809 |
| 10 | Depression | **rs4632195** | 18 | 53220378 | DCC | intron | 6.00E-18 | 34045744 | T | T | 0.50 | 0.48 | 72.07 | 2.03E-05 | 26.24 | 0.294 |
| 1 | Schizophrenia | rs11665242 | 18 | 53380757 | DCC | intron | 9.00E-10 | 33169155 | G | G | 0.40 | 0.16 | 105.20 | 2.05E-09 | -1.66 | 0.961 |
| 2 | Schizophrenia | rs1431196 | 18 | 53305732 | DCC | intron | 6.00E-11 | 31374203 | G | G | 0.41 | 0.16 | 104.70 | 2.19E-09 | 0.71 | 0.983 |
| 1 | Parkinson disease | rs8017172 | 14 | 55732330 | - | intergenic | 2.00E-07 | 28892059 | A | G | 0.45 | 0.30 | -132.30 | 3.36E-15 | 139.20 | 2.84E-07 |
| 2 | Parkinson disease | rs8012377 | 14 | 55724695 | LOC124903320 | downstream | 2.00E-10 | 38155330 | T | C | 0.47 | 0.28 | -127.80 | 3.14E-14 | 150.30 | 4.02E-08 |
| 3 | Parkinson disease | rs947211 | 1 | 205783537 | - | nc transcript<br>exon | 2.00E-12 | 19915576 | A | G | 0.28 | 0.49 | 77.38 | 5.19E-05 | -31.67 | 0.200 |
| 4 | Parkinson disease | rs6060983 | 20 | 31833121 | MYLK2 | intron | 6.00E-07 | 32201043 | C |  | 0.30 |  | -73.76 | 5.31E-05 |  |  |
| 5 | Parkinson disease | rs708726 | 1 | 205793011 | SLC41A1 | intron | 2.00E-08 | 37805635 | T | G | 0.28 | 0.45 | 76.35 | 6.22E-05 | -22.32 | 0.372 |
| 1 | ADHD | rs62098012 | 18 | 53335941 | DCC | intron | 7.00E-12 | 34446935 | T | T | 0.40 | 0.16 | 106.20 | 1.41E-09 | -0.30 | 0.993 |
| 2 | ADHD | rs17411548 | 18 | 53296573 | DCC | intron | 4.00E-14 | 35764056 | T | T | 0.40 | 0.17 | 104.30 | 2.43E-09 | -3.64 | 0.913 |
| 3 | ADHD | rs6508220 | 18 | 53304806 | DCC | intron | 7.00E-17 | 35764056 | A | A | 0.49 | 0.30 | 96.35 | 1.67E-08 | 4.34 | 0.869 |
| 4 | ADHD | rs9807529 | 18 | 53418531 | DCC | intron | 7.00E-11 | 35764056 | G | G | 0.42 | 0.18 | 96.55 | 2.52E-08 | -10.76 | 0.734 |
| 5 | ADHD | rs8083850 | 18 | 53180922 | DCC | intron | 7.00E-15 | 35764056 | G |  | 0.42 |  | -95.19 | 3.07E-08 |  |  |
| 6 | ADHD | rs11664788 | 18 | 53446816 | DCC | intron | 3.00E-13 | 35764056 | T | G | 0.46 | 0.36 | -90.30 | 1.24E-07 | -42.29 | 8.58E-02 |
| 7 | ADHD | rs11663156 | 18 | 53385388 | DCC | intron | 2.00E-10 | 35764056 | C | C | 0.32 | 0.23 | -88.31 | 6.82E-07 | 15.10 | 0.605 |
| 8 | ADHD | rs4513167 | 18 | 53162159 | DCC | intron | 2.00E-08 | 38565336 | T |  | 0.39 |  | -78.57 | 6.09E-06 |  |  |
| 9 | ADHD | rs965943 | 18 | 53331060 | DCC | intron | 1.00E-10 | 35764056 | G |  | 0.39 |  | -77.81 | 7.15E-06 |  |  |
| 10 | ADHD | rs10787738 | 10 | 117017860 | SHTN1 | intron | 3.00E-19 | 34446935 | T | T | 0.28 | 0.36 | 84.16 | 7.35E-06 | 18.86 | 0.449 |
| 11 | ADHD | **rs4632195** | 18 | 53220378 | DCC | intron | 4.00E-10 | 33479212 | T | T | 0.50 | 0.48 | 72.07 | 2.03E-05 | 26.24 | 0.294 |
| 1 | Bipolar | **rs11663393** | 18 | 53088362 | DCC | intron | 2.00E-10 | 31926635 | A | A | 0.45 | 0.32 | 72.85 | 1.72E-05 | 6.26 | 0.809 |
| 2 | Bipolar | **rs4632195** | 18 | 53220378 | DCC | intron | 4.00E-10 | 33479212 | T | T | 0.50 | 0.48 | 72.07 | 2.03E-05 | 26.24 | 0.294 |
| 1 | OCD | **rs8084351\*** | 18 | 53200189 | DCC | intron | 4.00E-12 | 31835028 | A | A | 0.49 | 0.42 | 66.38 | 8.92E-05 | -17.60 | 0.484 |

Table 5 presents the SNP associations with putamen volume at a threshold level of 0.05 in White and/or non-White participants of the ABCD dataset for the SNPs that were associated with at least three neuropsychiatric disorders, including depression ^48,49,61–65^, schizophrenia ^51,60,61,64–72^, Parkinson’s disease ^55^, ADHD ^57,58,60,61,64,71^, bipolar disorder ^48,60,61,64,65,68,73–77^, and OCD ^61^, as identified from the NHGRI GWAS catalog. A visualization of the full list of SNPs that shared an association between putamen volume at this threshold and two or more neuropsychiatric disorders is presented in Supplementary Figure 6, showing the sharing or overlap of significantly associated SNPs with putamen volume, depression, schizophrenia, Parkinson’s, ADHD, bipolar disorder, and OCD. There were 15, 9, 16, and 51 SNPs that shared with 5, 4, 3, and 2 neuropsychiatric disorders, respectively, and were also associated with putamen volume in in White and/or non-White participants of the ABCD dataset (Table 5 and Supp. Figure 6). These outcomes represent the shared genetic architecture of putamen volume and the neuropsychiatric disorders.

**TABLE 5:**
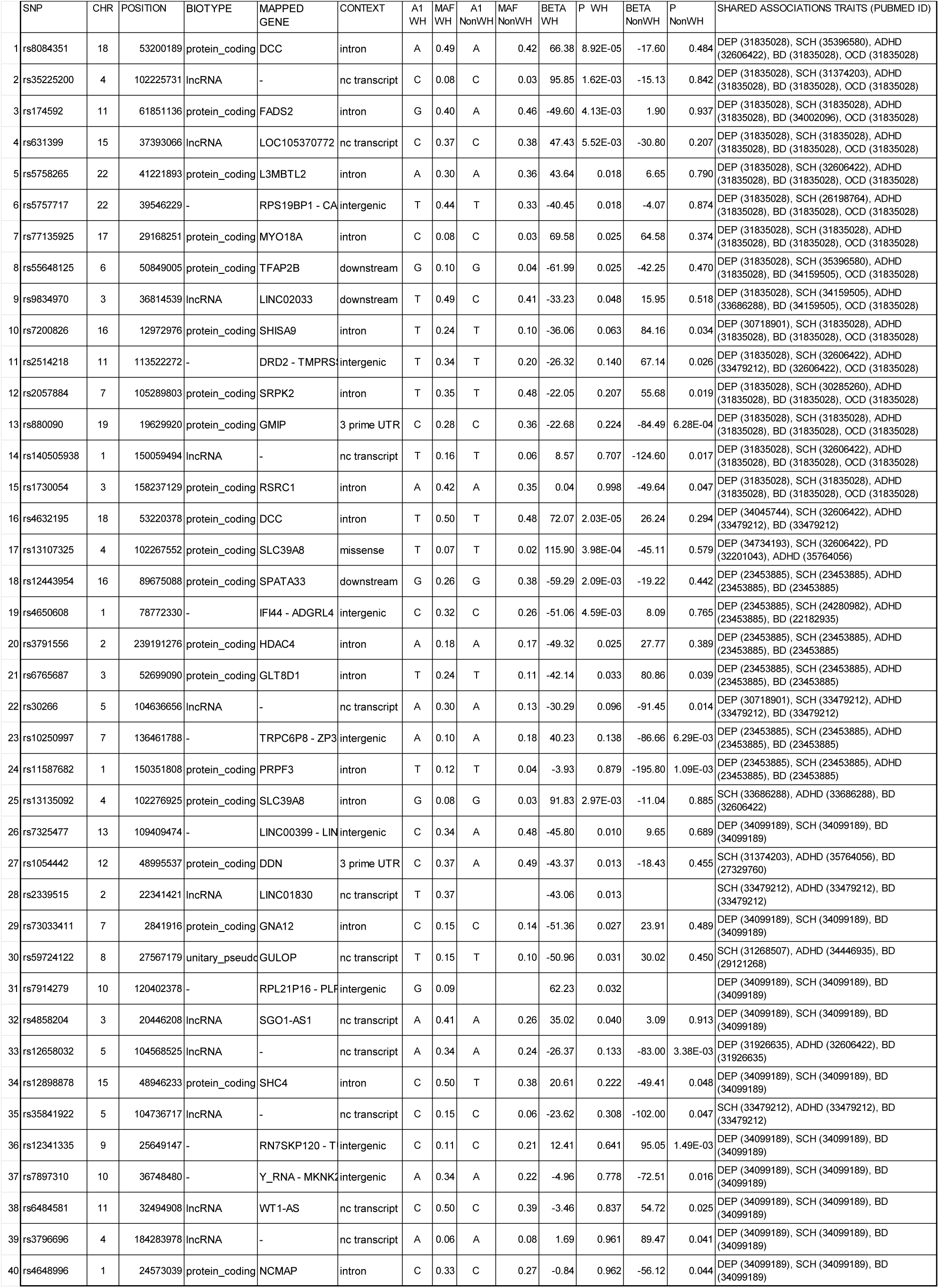
Shared genetic architecture of putamen volume and neuropsychiatric disorders, including depression (DEP), schizophrenia (SCH), Parkinson’s disease (PD), ADHD, bipolar disorder (BD), and OCD. The candidate SNP associations with putamen volume were assessed at the 0.05 significance threshold in ABCD White and/or non-White participants. The listed SNP shared associations with at least <u>three</u> neuropsychiatric disorders, as identified from NHGRI GWAS catalog, in addition to their associations with putamen volume in ABCD dataset.

The overlap between newly identified putamen volume GWAS SNPs, replicated candidate SNPs, and SNPs associated with neuropsychiatric disorders from prior GWASs is also presented in the last column of Table 1.

## DISCUSSION

In this study, we successfully evaluated the genetic main effect on putamen volume using GWAS in ABCD White participants and replicated most of the significant SNP associations in non-White participants. We performed pathway enrichment scoring analysis using the GWAS results and successfully replicated the associations of candidate SNPs from prior genetic studies on putamen volumes in the ABCD White and/or non-White participants. In addition, we assessed the shared genetic associations between putamen volume and multiple neuropsychiatric disorders, i.e., depression, schizophrenia, Parkinson’s disease, ADHD, bipolar disorder, and OCD. The most notable contributions were the following: 1) We identified 199 SNPs with genome-wide significant SNP-putamen volume associations in ABCD White participants. 2) Of these, more than a dozen of the most significant SNP associations were replicated in non-White participants, including five regulatory region SNPs (i.e., rs8017172, rs1953350, rs1953351, rs1959089, and rs1953352). 3) The association of about two dozen SNPs with putamen volumes from prior studies ^17,19,20,42–44^ were replicated in our analysis of the ABCD study dataset. 4) Despite an overlap in the SNP associations, we discovered that there is significant genetic heterogeneity between White and non-White participants in context of putamen-linked SNPs. 5) We identified a key pathway REACTOME_DSCAM_INTERACTIONS (https://www.reactome.org/content/detail/R-HSA-376172) linked with putamen volume that involves the *DSCAM* gene, netrin-1 protein, and *DCC* gene, depending on the cellular and developmental context ^39,41^. 6) We identified shared genetic associations with putamen volume and half-dozen neuropsychiatric disorders (depression, schizophrenia, Parkinson’s disease, ADHD, bipolar disorder, and OCD).

The most notable genes identified in our study were the *DCC* and *DSCAM* genes. Published literature suggests that *DCC* is a transmembrane protein with immunoglobulin and fibronectin domains ^34^, and the *DCC* axon guidance receptor and its ligands, the netrins, are crucial for guiding axonal projections toward the ventral midline in the developing nervous system ^35,36^. Netrins act as chemoattractants or chemorepellents for axons ^78^. Netrin-1 binding to *DCC* causes chemoattraction, while interaction with *UNC5* leads to chemorepulsion ^79^. *DCC* and netrin-1 are essential for commissural axon guidance and neuronal migration ^34,37^. *DSCAM*s, i.e., Down syndrome cell adhesion molecules, immunoglobulin-like transmembrane proteins with fibronectin III domains, were identified on chromosome 21 and regulate neuronal wiring in humans and other vertebrates ^38^. During nervous system development, spinal commissural axons are guided by netrin-1, which activates the *DCC* receptor, while *DSCAM* binding to netrin-1 is essential for axon guidance, acting independently or with *DCC* ^39^. Local translation of synaptic mRNAs, crucial for CNS development, is impaired in intellectual disabilities like Down syndrome, where *DSCAM* may be locally translated in dendrites and axonal growth cones of hippocampal neurons ^40^.

More than a dozen of the most significant SNP-putamen volume associations from White participants were replicated in non-White participants. We also successfully replicated SNP-putamen volume associations from prior studies in White and/or non-White participants, but we observed reduced p-value significance in these replications. This could be partly due to differences in participants size, population structure, genetic heterogeneity, or context-specific effects across populations. Overall, our study underscores the importance of replication in validating genetic associations and highlights the complexity of genetic influences across different groups.

Our findings of SNP-putamen volume associations are consistent with the high heritability (i.e., 71–79% ^17^) of putamen volume. However, epigenetic influences may overshadow the genetics influences ^80^ particularly in the presence of stressors ^81 82,83^. Observing relatively fewer replications in non-White participants, which also include Black participants among others, may be partly due to the fact that a substantial body of research indicates that non-White individuals, particularly Black people, are exposed to higher levels of stress compared to other racial groups ^84^. For example, factors such as prenatal stress ^85^, adverse childhood experiences (ACEs) ^86^, and parental depression ^87^ may give rise to epigenetic changes like DNA methylation ^88^, which in turn may affect putamen volume. Particularly, ACEs have a profound impact on mental and physical health, disrupting brain development in regions related to stress regulation, including the prefrontal cortex, hippocampus, amygdala, and putamen ^89^, contributing to mental health disorders ^90^. Parental depression, particularly maternal, is also linked with reduced putamen volume in children ^87,91^. Further studies are needed to evaluate the influence of early-life stressors and epigenetics on putamen volume and associated neuropsychiatric disorders.

In conclusion, we identified a comprehensive list of genome-wide significant SNPs associated with putamen volume, discovering (i) robust and replicable regulatory SNP associations, (ii) genetic heterogeneity in putamen-linked SNPs, (iii) biological pathways involving genes associated to putamen volume, and (iv) genetic associations that are shared between putamen volume and several neuropsychiatric disorders, including depression, schizophrenia, Parkinson’s disease, ADHD, bipolar disorder, and OCD. Our findings provide a deeper understanding of the genetic architecture and cross-population differences associated with putamen volume, which may help with the advancement of precision medicine.

## DATA AVAILABILITY

The source data for the Adolescent Brain and Cognitive Development study, which was used for the secondary analysis in this study, was publicly available prior to the initiation of the study. The dataset can be accessed through authorized data access from the NIH Brain Development Cohorts (NBDC) Data Hub (https://www.nbdc-datahub.org/abcd-study).

## ACKNOWLEDGEMENT

This study was partially supported by the National Institute of Mental Health (NIMH) grant 1R01MH133313-01A1. We also thank the investigators, staff, and participants of the ABCD study for their valuable contributions in generating and making the study dataset publicly available.

