## SUPPLEMENTARY FIGURES for "Identification of Comprehensive Genetic Factors, Pathways, and Shared Genetic Architecture of Putamen Volume in Adolescent Cohort"

*Correspondence:

Abanish Singh, Ph.D.

Supplementary Figure 1: Ven diagram of race-wise ABCD study sample size.

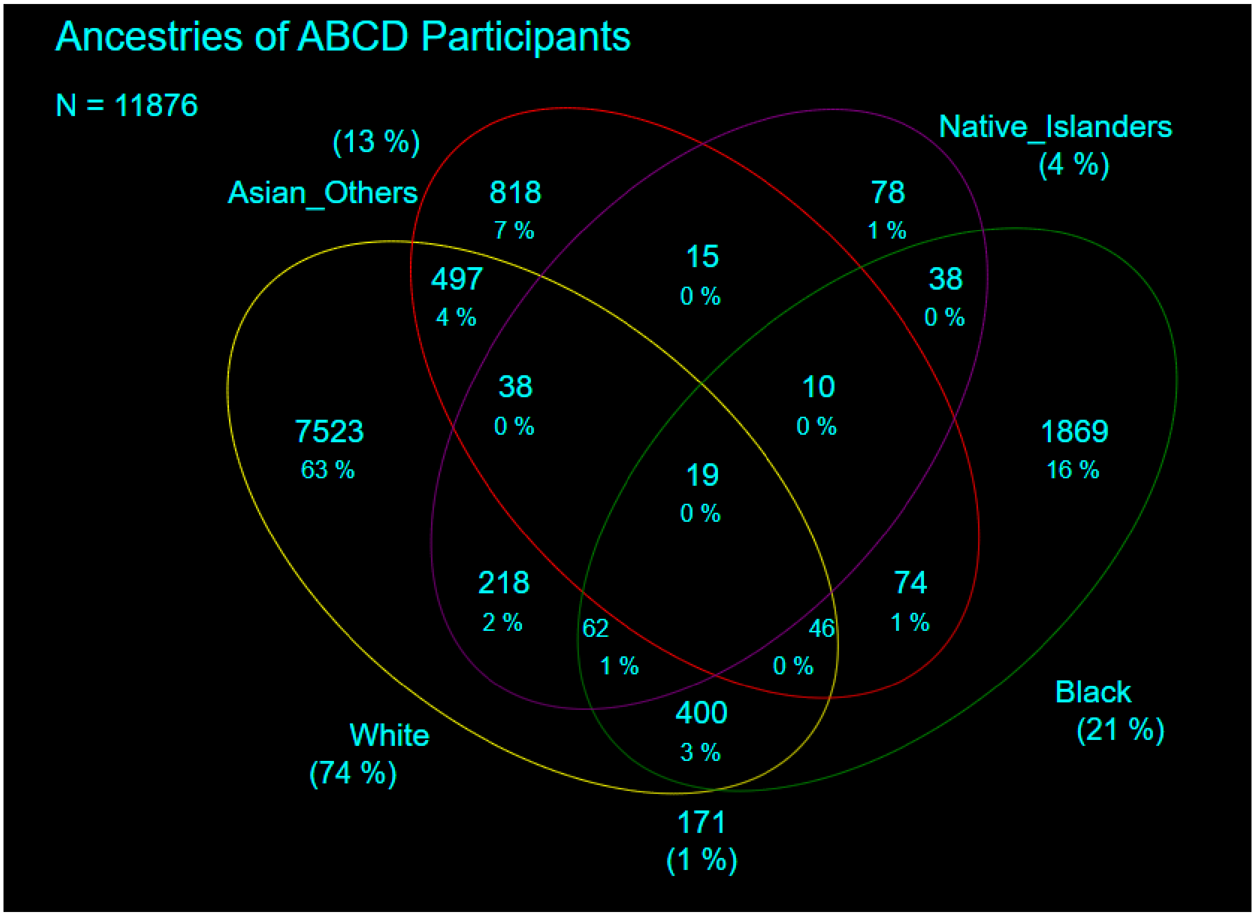

Supplementary Figure 2: Scree plot for principal component analysis (PCA) of White participants' genotypes.

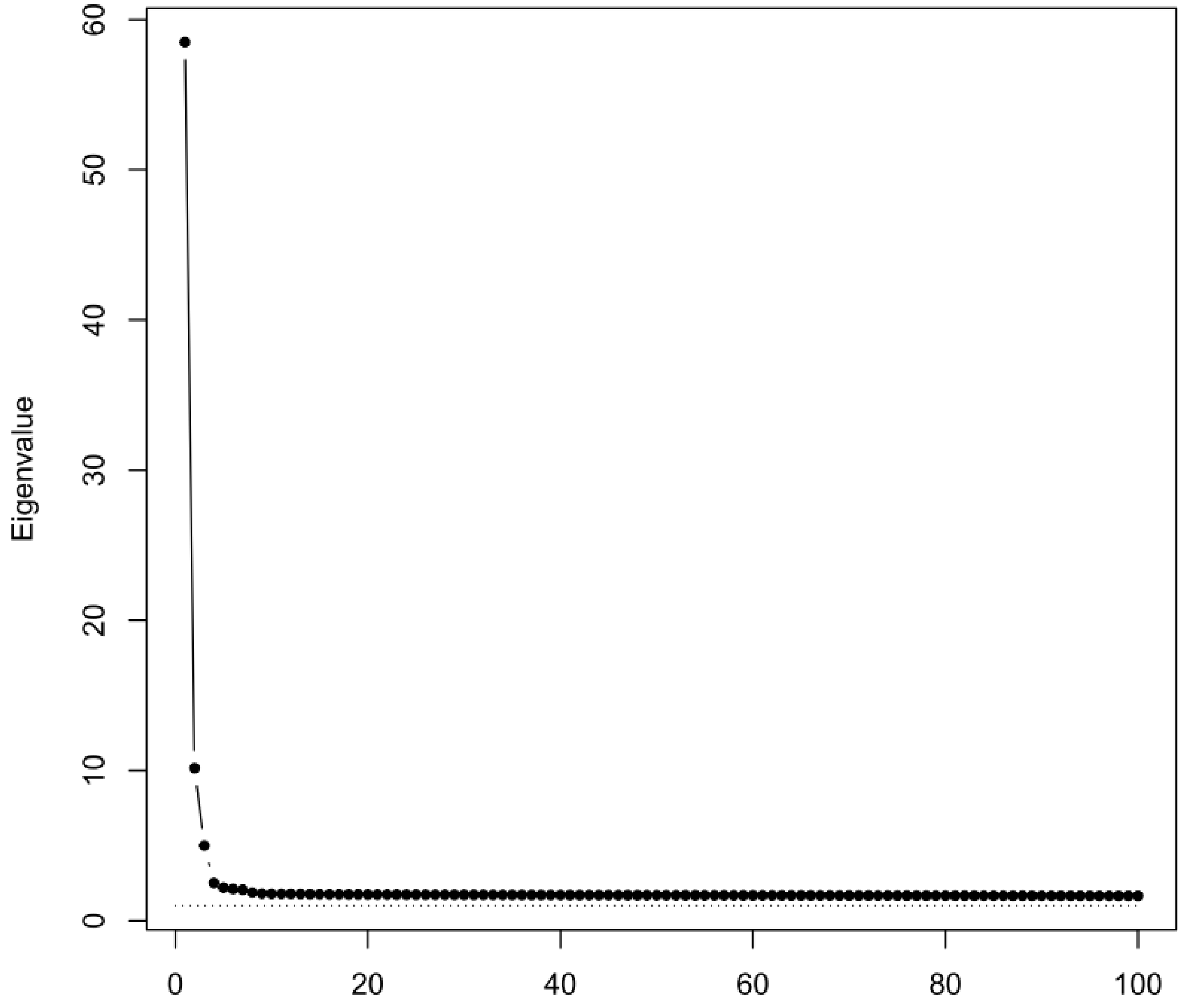

Supplementary Figure 3: Scree plot for principal component analysis (PCA) of Non-White participants' genotypes.

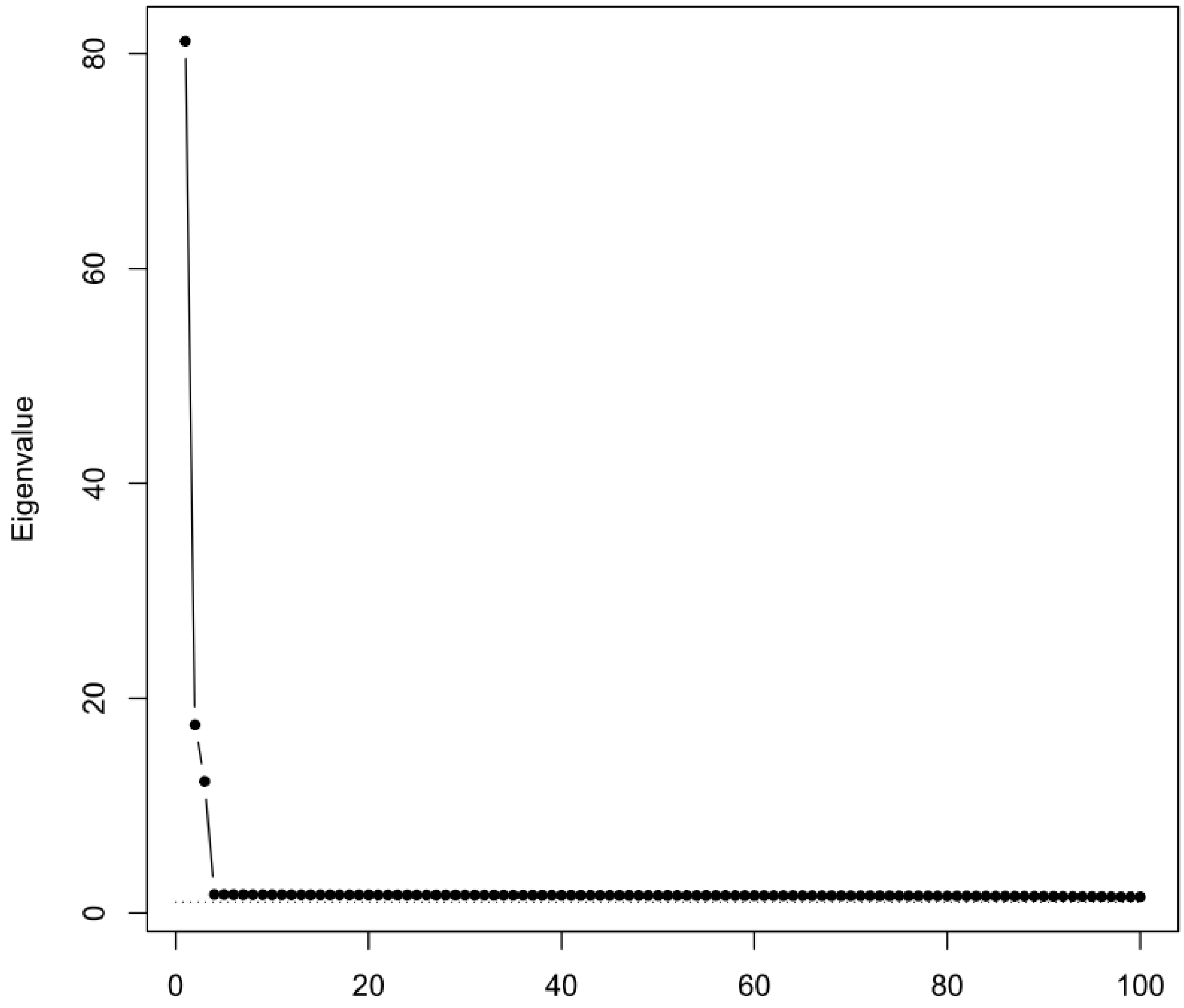

Supplementary Figure 4: Distribution of putamen volume in White participants.

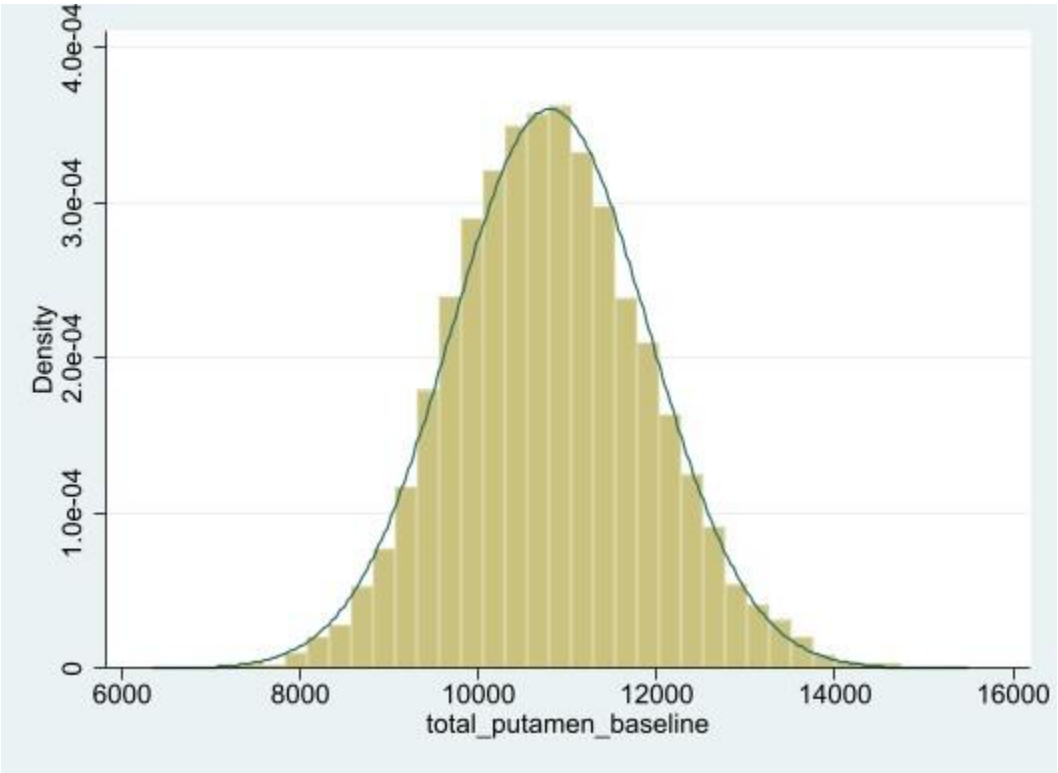

Supplementary Figure 5: Distribution of putamen volume in non-White participants.

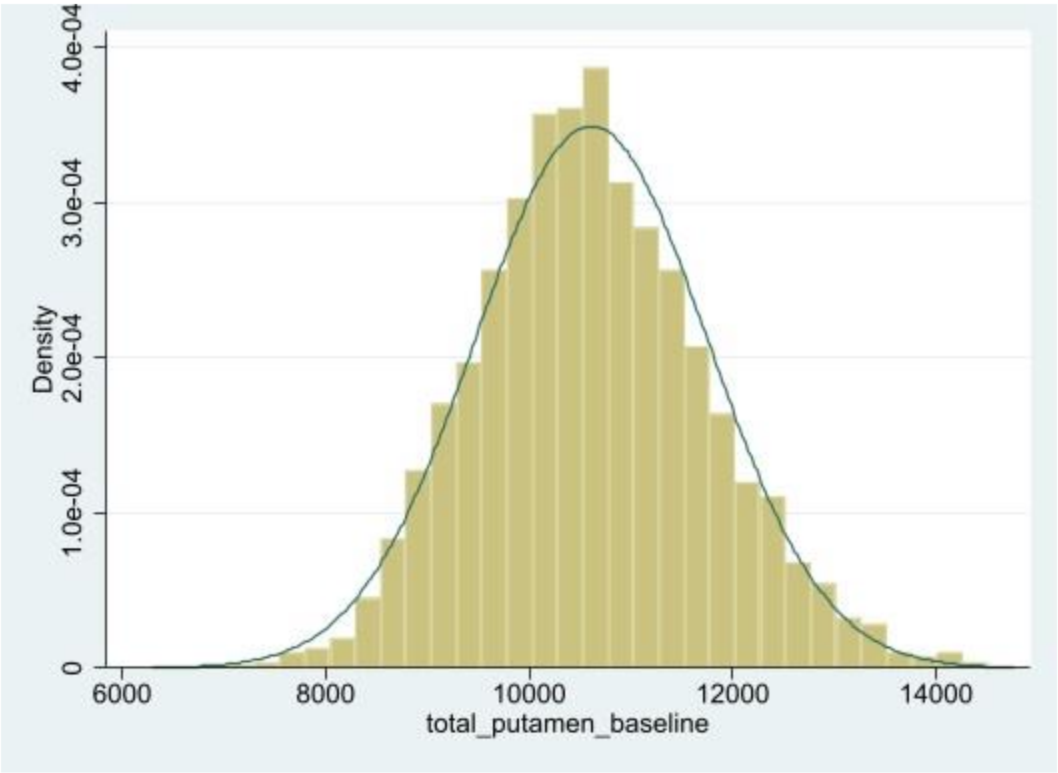

Supplementary Figure 6: A circular plot showing the overlap of significantly associated SNPs (represented by red dots) with putamen, depression, schizophrenia, Parkinson's, ADHD, bipolar disorder, and OCD, displayed in order from the outermost to the innermost tracks, respectively.

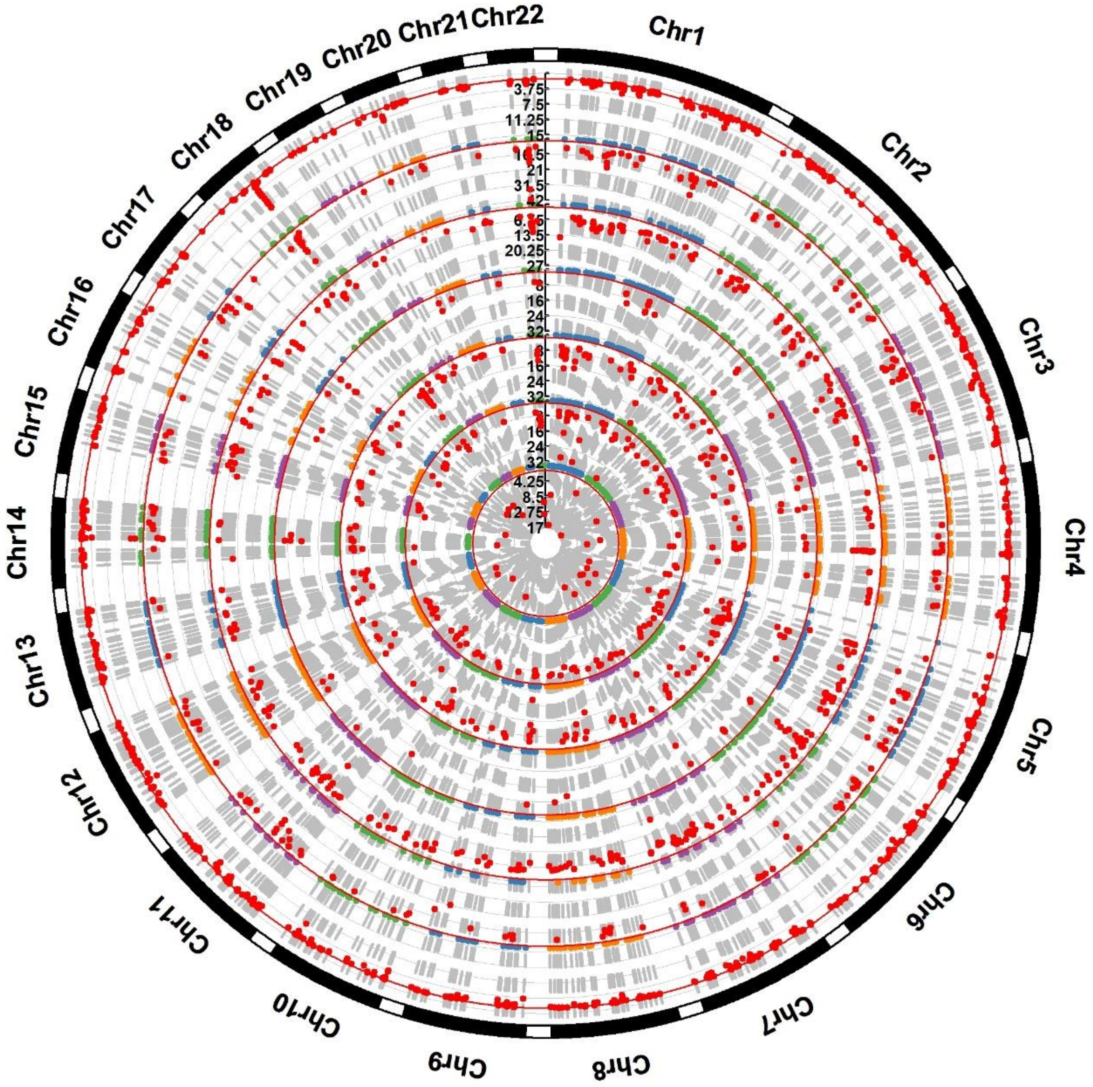
